# Survey of COVID-19 associated symptoms and reported deaths in an urban community in Kano, Nigeria

**DOI:** 10.1101/2021.06.14.21258236

**Authors:** Disha Shahani, Zayyad Sani Farouq, Hadiza Galadima, Ashna Khare, Nirmal Ravi

**Author notes:** Corresponding Author: Nirmal Ravi^1^, 4-6 Independence Rd, Kano, Kano, Nigeria.

## Abstract

**Background:** Nigeria reported the first case of COVID-19 on February 27, 2020. By June of 2020, many people reported experiencing mild COVID-19 associated symptoms, yet did not get tested due to inaccessible testing and insufficient knowledge of the disease. There were media stories quoting grave diggers in Kano who reported high burial rates during this time.

**Methods:** In order to draw more data on COVID-19 cases during this time period, we conducted a cross-sectional symptom survey in Kano, surveying 291 adults. Participants were asked to report demographic characteristics, past COVID-19 testing and symptoms, and community deaths. To assess associations between COVID-19 associated symptoms and socio-demographic characteristics, bivariate analyses using Chi-square tests were performed. A logistic regression assessing the association between any reported symptoms and the kind of work (indoor/outdoor) was done while adjusting for age, gender and education level.

**Results:** Half of the respondents reported at least one symptom associated with COVID-19; the three most common symptoms were loss of appetite, cough, and fever. There was a statistically significant relationship between age group of the respondent and presence of COVID-19 associated symptoms. Gender or level of education did not have statistically significant association with COVID-19 associated symptoms among the respondents. People with outdoor occupations such as trading and hawking were more than twice as likely to report COVID-19 associated symptoms compared to those who were unemployed. Just under half of the respondents reported knowing someone who died in their community, with unexplained causes attributed to two-thirds of these cases. Our study found evidence of COVID-19 associated symptoms especially among the older population and unexplained deaths in Kano. Lack of confirmatory laboratory tests and absence of baseline vital statistics precluded us from finding definitive evidence for or against COVID-19 infection and associated mortality.

## Introduction

In December 2019, Wuhan City, Hubei, China experienced an outbreak of coronavirus disease (COVID-19), caused by SARS-CoV-2. The disease rapidly evolved into a global emergency, causing the World Health Organization (WHO) to declare a Public Health Emergency of International Concern in January 2020 (Harapan, Itoh et al. 2020). As of March 2021, there have been a reported 122 million cases, with over 2.7 million deaths worldwide. (Dong Lancet Online Dashboard).

SARS-CoV-2 spreads from an infected person to another through respiratory droplets released during activities such as coughing, sneezing, talking and breathing. A meta-analysis of 18 studies concluded that the average incubation period of SARS-CoV-2 was 5.7 days and that the incubation period varied between countries (Wassie, Azene et al. 2020). The signs and symptoms vary from fever, cough and fatigue to headache, diarrhea, myalgia, loss of sensation of smell, hemoptysis, dyspnea, and lymphopenia (Huang, Wang et al. 2020). A study of 1480 participants with flu-like symptoms found that loss of smell and taste were positively associated with diagnosis of COVID-19 whereas, sore throat was strongly associated with negative tests (Yan, Faraji et al. 2020). Despite about a third of participants in a study reporting loss of smell/taste as their initial symptom, they were less likely to get tested for COVID-19 unless they also had other accompanying symptoms (Coelho, Kons et al. 2020). Prevalence of gastrointestinal symptoms has been studied sparsely, and a meta-analysis reported nausea or vomiting, diarrhea and loss of appetite as the most common pooled gastro-intestinal symptoms (Mao, Qiu et al. 2020, Schmulson, Davalos et al. 2020)

The outbreak that began in China quickly spread to other parts of the world. Africa began preparing for the pandemic as early as January 2, 2020, implementing strict screening procedures at airports, and surveying those with a recent travel history to China or Europe (Massinga Loembe, Tshangela et al. 2020). Statistical models pointed to Egypt, Algeria, and South Africa at the greatest risk of having the virus spread to Africa, and Nigeria at moderate risk of importation of the disease (Gilbert, Pullano et al. 2020). Early models of COVID-19 spread in Nigeria estimated that the disease could be controlled by May 2020 (Zhao, Li et al. 2020).

Nigeria reported the first laboratory confirmed case of COVID-19 on February 27, 2020 (Elimian, Ochu et al. 2020). As of July 2, 2020, Nigeria was only able to test 2,500 COVID-19 samples everyday due to the lack of testing availability, laboratory equipment, and scientists. Nigeria had tested 728,128 samples out of a population of 200 million by November 2020 (NCDC, 2020). This is in sharp contrast to South Africa, a wealthier country with greater resources, that had tested over 1.6 million people of its total 58 million population (Dixit S, 2020). This study highlights the gap between wealthy and poor countries; the latter having significantly fewer resources to deal with the global pandemic. Despite reports that South Africa has been hit the hardest in Africa by COVID-19, this is not a reliable indication as to whether or not it truly has the greatest number of cases as demonstrated by the fact that the country has had more accessibility to testing (Gaye, Khoury et al. 2020).

Serosurveillance has been a leading form of measuring prevalence of COVID-19 at a population level. The National Centre for Immunisation Research and Surveillance (NCIRS) has been testing 7,000-13,000 blood samples for antibodies of SARS-CoV-2 in Australia (Serosurveillance NCIRS, 2020). Scotland conducted a surveillance of SARS-CoV-2 IgG antibodies in residual blood samples from clinical laboratories that showed 4.3% seroprevalence with no variation by age, sex or geographic location (Dickson, Palmateer et al. 2021). Such studies are supposed to assist in examining immunity trends, evaluating vaccination impact, predicting outbreaks, modeling disease, and affecting immunization policy (Serosurveillance NCIRS, 2020). Nevertheless, such sero-surveys are not generalizable to the worldwide population and may only be applicable to certain regions and/or cohorts. Most of these studies are being conducted in wealthier countries which often have better resources to conduct serosurveillance. In addition to other reasons, Europe used serosurveillance to document school outbreaks, dividing students by age group and subsequently conducting contact tracing and other analysis to deduce symptomatic information (Otte Im Kampe, Lehfeld et al. 2020). A small serosurveillance study conducted in June in Niger state in Nigeria found 25% seropositivity level (Majiya, Aliyu-Paiko et al. 2020).

In June 2020 during the data gathering phase of this study, COVID-19 testing in Kano, Nigeria was made available only for patients meeting the Nigeria Center for Disease Control’s (NCDC) case definition for COVID-19. A suspected COVID-19 case according to the NCDC was anyone with cough and/or fever with 1 or more other symptoms [NCDC Community Case Definitions for Coronavirus Disease, 2020]. However, as previously discussed, although fever was positively associated with tests, chemosensory dysfunctions were extremely common among COVID-19 patients and often preceded fever as a presenting symptom (Mercante, Ferreli et al. 2020, Yan, Faraji et al. 2020). With the Nigeria Center for Disease Control’s case definition, potential COVID-19 cases were missed if individuals with only chemosensory loss were less likely to be tested as was seen in the United States (Coelho, Kons et al. 2020).

There were anecdotal reports in Nigeria during June 2020 of many people experiencing symptoms such as loss of smell but no other symptoms. Most people with symptoms did not get tested for COVID-19, either because they had insufficient knowledge of the disease or because testing was not available or accessible. A study done in Ibadan, Nigeria showed strong denial of COVID-19 and distrust of government efforts to tackle it (Ilesanmi and Afolabi 2020). Media also quoted gravediggers in Kano reporting unusually high burial rates during this period (The Economist, 2020). We did a community survey in June 2020 in an urban community in Kano to document the presence of COVID-19 associated symptoms and deaths. We also wanted to test for associations between demographic variables and presence of COVID-19 associated symptoms.

## Materials & Methods

A cross-sectional study was conducted in Kano, Nigeria, comprising 291 adult (at least 18 years of age) respondents from Nasarawa Local Government Area. Ethics approval was obtained from Kano state government ministry of health (Approval number MOH/Off/797/T.I/2052). Informed verbal consent was obtained from all participants before data was collected from them. The eventual sample population who consented to participate in the study was 286.

### Variables and measurements

The survey asked for the following self-reported data by the participants: demographic characteristics (age, gender, occupational status, type of work, religion, marital status), visit to healthcare workers, testing for COVID-19, and having experienced any COVID-19 associated symptoms. The survey also consisted of questions about any known deaths in the community, cause of death and symptoms experienced.

### COVID-19 associated symptoms and test

Participants were asked if they experienced any of the symptoms from a list during the month of Ramadan (June 2020). Each respondent’s symptoms were grouped categorically; none, cough, fever, shortness of breath, sore throat, fatigue, loss of appetite, loss of smell (anosmia) and myalgia. Symptoms were also dichotomously coded as having expressed none or at least one symptom. Participants were asked if they got tested for COVID-19 during the month of Ramadan.

### Employment status and type

The questionnaire asked participants to report employment status (Yes/No) and occupation (self-reported description). Amongst those employed, occupation was further categorized as indoor and outdoor work.

Individuals categorized as outdoor work included those working as guards, police, farming and those involved in poultry work, fruit and vegetable sellers, traders and shop owners, mechanics, welders, carpenters, construction workers and shoemakers.

Individuals categorized as indoor work were those involved in businesses, accounting, banking, pharmacy, laundry, library assistance, painting, tailoring, company or factory workers, academics (teachers, lecturers, scholars and hisbah workers), nannies, drivers, and stylists (henna designer, hair stylist, salon worker, cosmetics).

### Education level

Amongst those who claimed to have attended school (Yes/No), education was categorized as adult literacy, primary level, secondary level, tertiary level and Quranic studies.

### Age

Age was described as a continuous variable as well as a categorical variable.

### Known deaths in the community

Participants were queried on the death of any known individuals in their communities during the month of Ramadan, their relation to them (close family member, close neighbor, extended family member, casual acquaintance or a work colleague), cause of their death and if they had any COVID-19 associated symptoms.

### Statistical Analysis

Statistical analyses were conducted using STATA MP 13 (Lakeway Drive College Station, Texas). Descriptive and inferential statistical methods were used to summarize the data. Categorical variables were presented as frequencies and proportions, whereas continuous variables were presented as median and interquartile ranges. To assess associations between COVID-19 associated symptoms and socio-demographic characteristics, bivariate analyses using Chi-square tests were performed. A logistic regression assessing the association between any reported symptoms and the kind of work (indoor/outdoor) was done while adjusting for age, gender and education level. An alpha level of 0.05 was used to determine statistical significance.

## Results

### Social and demographic characteristics

Socio-demographic characteristics of the respondents are presented in table 1. The median age of the respondents was 36 years with an interquartile range (IQR) of 26 – 48 years. More than half of the sample population were female (53.85%, n=154) and about one-third were married (33.57%, n=96). Nearly all (96%, n=265) the respondents had at least some formal education. Less than one-third (29.37%, n = 84) of the respondents had attained education at the tertiary level, followed by secondary education (24.48%, n = 70) and primary education (13.64%, n=39). One-fifth of the respondents had only attended Quranic school. More than half the respondents (58.04%, n= 166) reported being gainfully employed. About a quarter (26.22%, n=75) of the working population worked outdoors.

**Table 1.** Socio-demographic characteristics of survey population from Kano, Nigeria

| Characteristics | n | % |
| --- | --- | --- |
| **Age group** |  |  |
| 18-34 | 121 | 42.31 |
| 35-49 | 102 | 35.66 |
| 50-64 | 54 | 18.88 |
| 65+ | 9 | 3.15 |
| **Gender** |  |  |
| Female | 154 | 53.85 |
| Male | 132 | 46.15 |
| **Religion** |  |  |
| Christianity | 30 | 10.49 |
| Islam | 256 | 89.51 |
| **Formal education** |  |  |
| No | 21 | 7.34 |
| Yes | 265 | 92.66 |
| **Level of education** |  |  |
| None | 21 | 7.34 |
| Primary | 39 | 13.64 |
| Secondary | 70 | 24.48 |
| Tertiary | 84 | 29.37 |
| Adult education class | 13 | 4.55 |
| Quranic | 59 | 20.63 |
| **Marital Status** |  |  |
| Married | 96 | 33.57 |
| Not married | 190 | 66.43 |
| **Employed** |  |  |
| No | 123 | 43.01 |
| Yes | 163 | 56.99 |
| **Type of occupation** |  |  |
| No occupation | 123 | 43.01 |
| Outdoor | 75 | 26.22 |
| Indoor | 88 | 30.77 |

About half (46.50%, n=133) of the respondents reported COVID-19 associated symptoms as seen in table 2. Only 1.40% of the survey participants had reported being tested for COVID-19 whereas a quarter of the participants (25.52%, n=73) reported visiting a healthcare worker during the month of June.

**Table 2.** COVID-19 indicators

| **COVID-19 questions** | **n** | **%** |
| --- | --- | --- |
| **Reported at least one COVID-19 associated symptom** |  |  |
| No | 153 | 53.5 |
| Yes | 133 | 46.5 |
| **Tested for COVID-19** |  |  |
| No | 282 | 98.6 |
| Yes | 4 | 1.4 |
| **Visited HCW** |  |  |
| No | 213 | 74.48 |
| Yes | 73 | 25.52 |

### COVID-19 associated symptoms among respondents

The distribution of COVID-19 associated symptoms among the different age groups can be seen in figure 1. Roughly half (53.50%, n=153) of all respondents did not report any symptoms. The three most frequent symptoms reported among all respondents were loss of appetite (17.48%, n=50), cough (17.48%, n=50) and fever (17.13%, n=49). Shortness of breath was the least reported symptom among all respondents (3.5%, n=10). Anosmia was reported by 6.64% of respondents.

**Figure 1.**
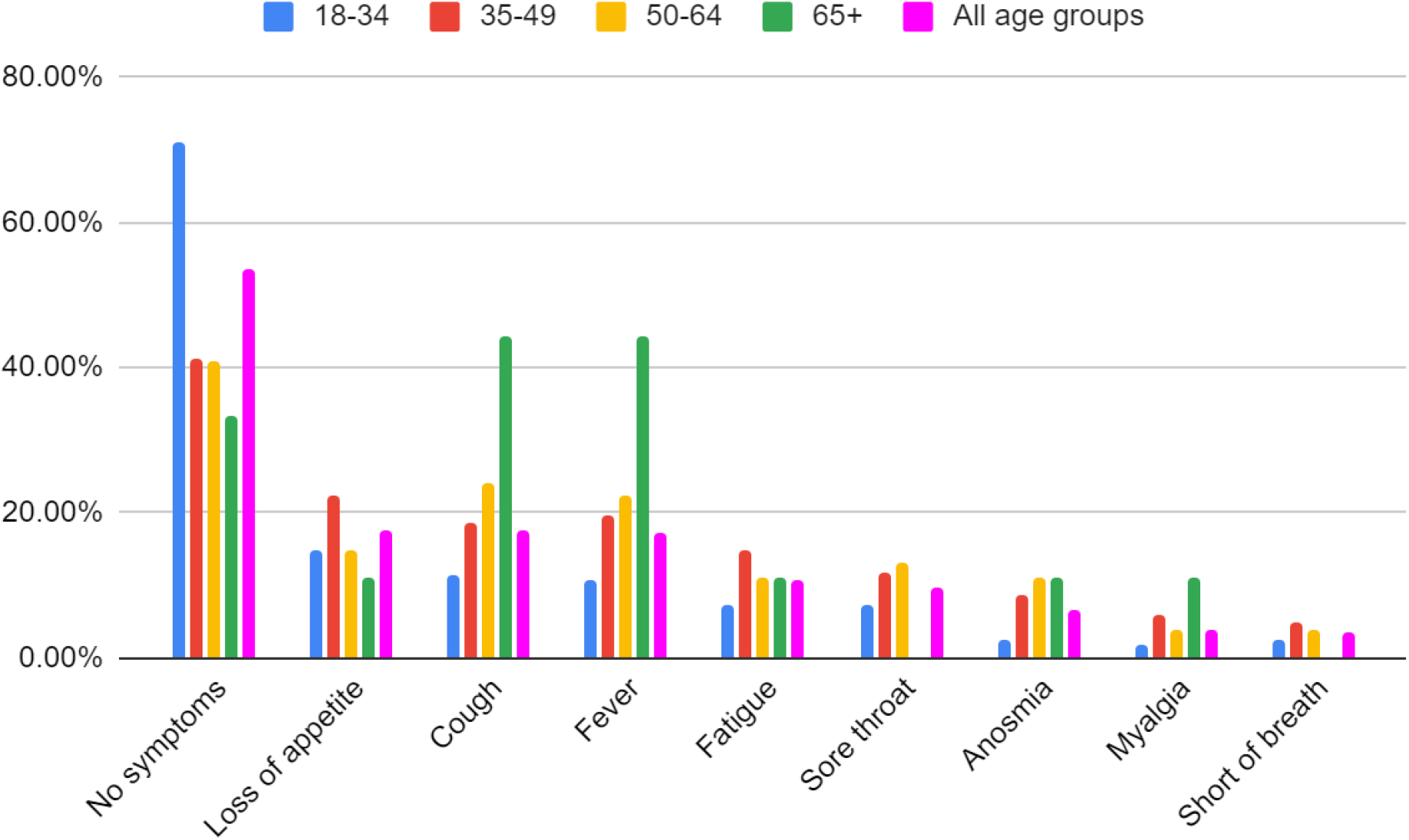
Distribution of symptoms by age groups among the respondents

The distribution of symptoms varied depending on the age group. Younger age groups were more likely to report no symptoms compared to older age groups. The proportion of respondents reporting no symptoms was highest in the 18-34 age groups. Symptoms of cough and fever were the highest in the 65+ age group followed by 50-64 age group. There was a statistically significant relationship between age group and the presence of COVID-19 associated symptoms (p=0.000).

Additionally, figure 2 shows the distribution of COVID-19 associated symptoms by gender. Slightly fewer than half the female respondents (49.35%, n=76) reported no symptoms while more than half the male respondents reported no symptoms. All the COVID-19 associated symptoms except myalgia were more common among females than males. However, there was no statistically significant relationship between gender and the presence of COVID-19 associated symptoms (p=0.165).

**Figure 2.**
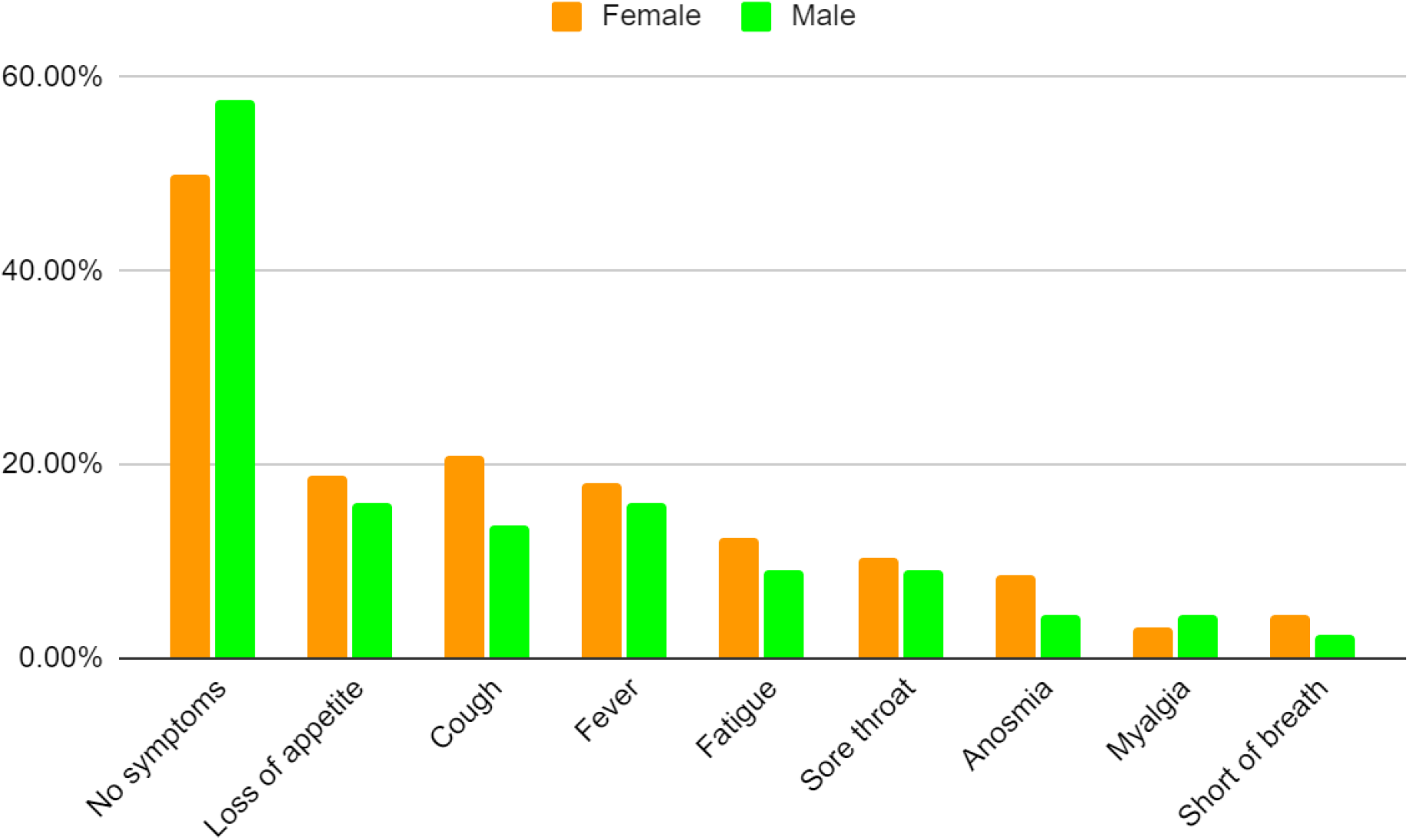
Distribution of symptoms by gender

Table 3 shows the adjusted odds ratio of reporting COVID-19 associated symptoms and sociodemographic characteristics. The odds of experiencing symptoms was strongly associated with increasing age (aOR 1.04 95% CI: 1.02-1.06). When occupation was categorized as indoor or outdoor work, it was found that those with symptoms had 2.18 times the odds (95% CI: 1.13 - 4.20) and 1.71 times the odds (95% CI: 0.92 – 3.20) of reporting working outdoors and indoors respectively, than those who did not report any symptoms. The association between indoor work and experiencing symptoms was not statistically significant (p=0.09). We also found that men had 0.72 times the odds (95% CI: 0.42-1.25) of women to report COVID-19 associated symptoms. This was not statistically significant (p=0.24). There were decreasing odds of reporting COVID-19 symptoms with increasing levels of education when adjusted for gender, type of work and age. The associations between primary, secondary, tertiary, adult, and quranic education and reporting COVID-19 associated symptoms were not statistically significant (p=0.77, p=0.95, p=0.33, p=0.93, p=0.32 respectively).

**Table 3:**
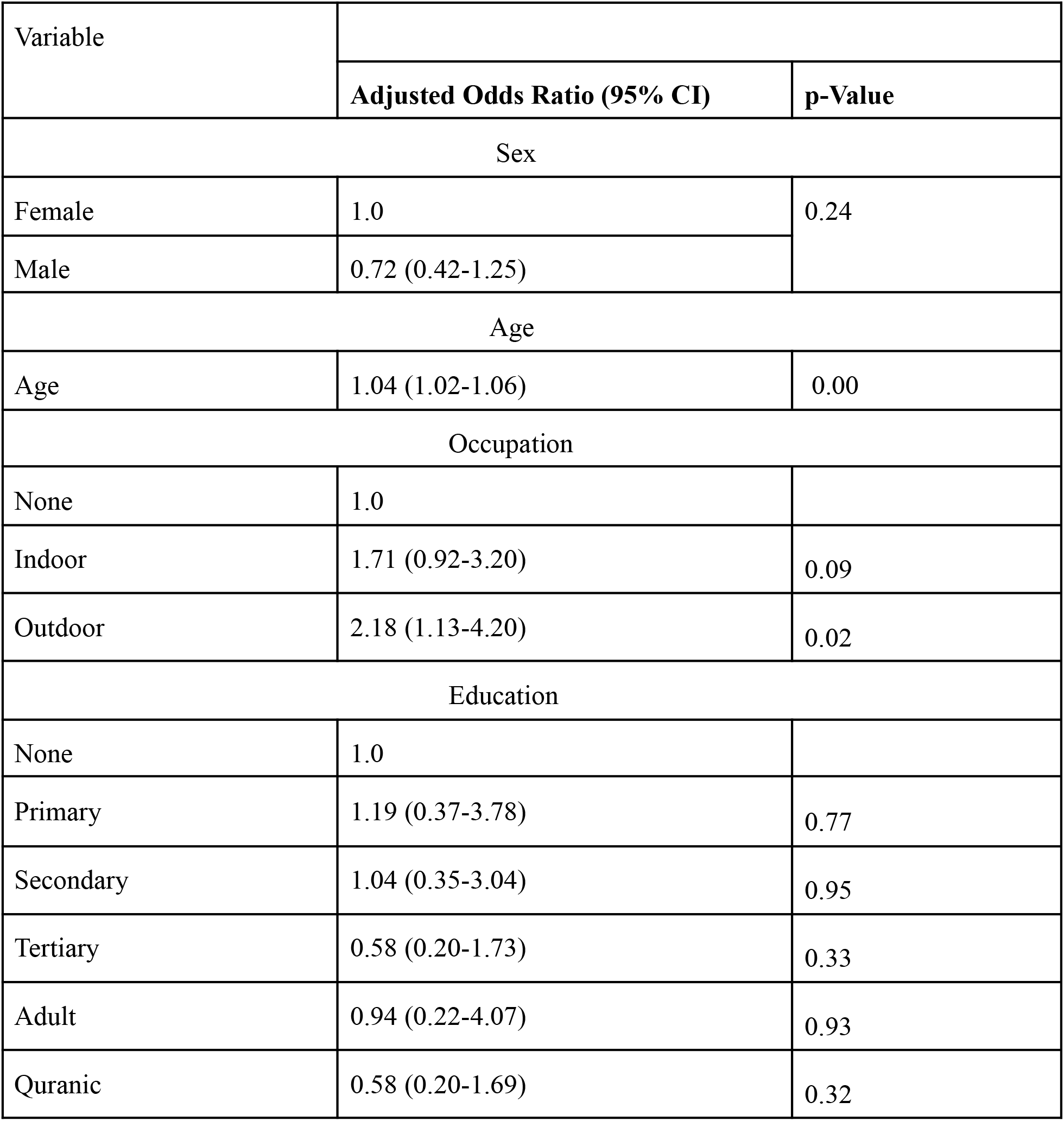
**Adjusted Odds Ratio of reporting COVID-19 associated symptoms and socio-demographic characteristics**

### Deaths in the community

Figure 3 shows the distribution of COVID-19 associated symptoms among the deceased as reported by the respondents. The three most frequent symptoms reported to be experienced by the deceased were fever (28.57%, n=34), fatigue (13.45%, n=16) and loss of appetite (11.76%, n=14).

**Figure 3.**
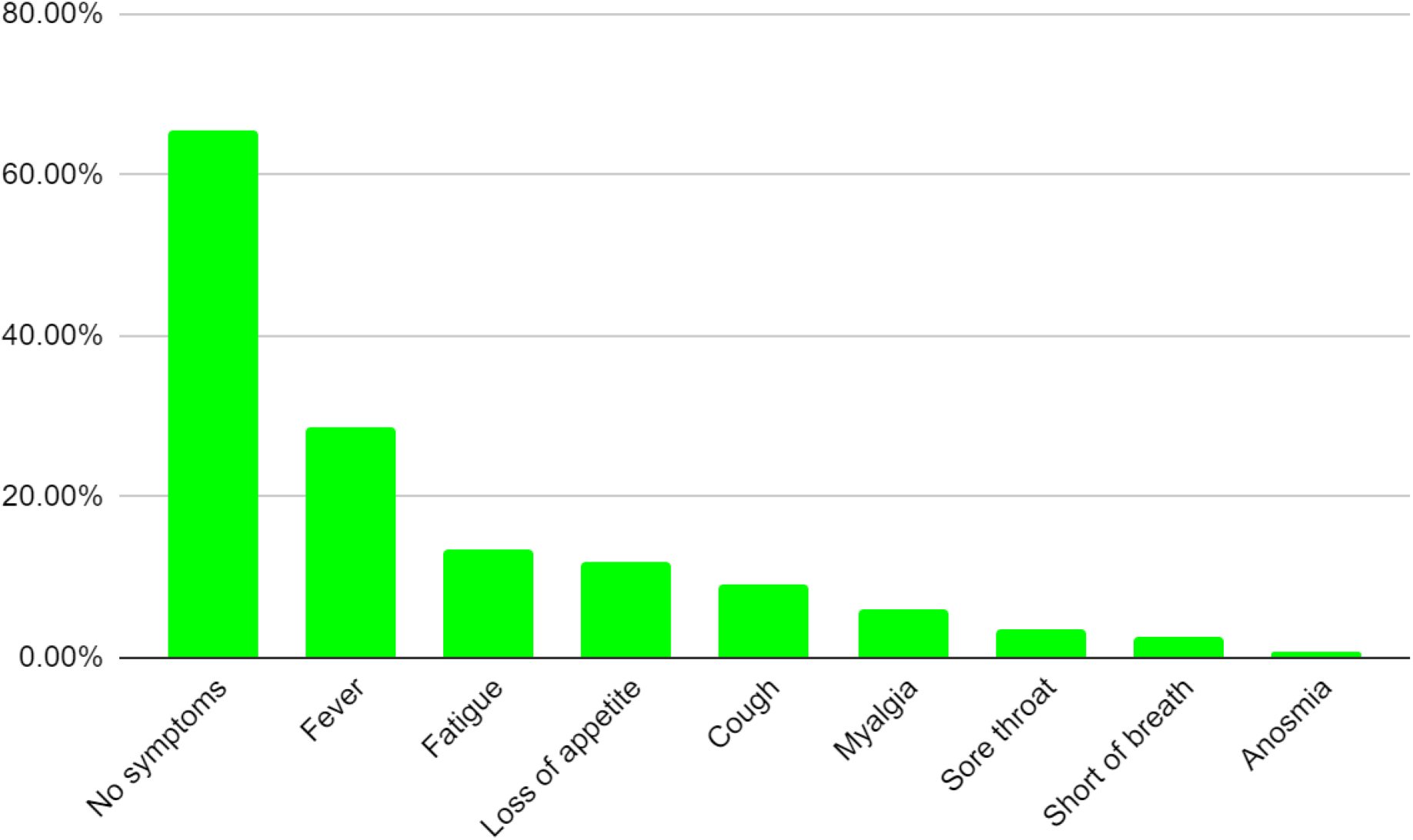
Distribution of symptoms among deceased

Figure 4 shows the distribution of community deaths reported by the respondents and their relationship to the deceased individuals. Slightly more than half the respondents (58.39%, n= 167) reported that they did not know of any deaths in their communities while 15% (n=43) reported the death of a neighbor, 9.44% (n=27) reported the death of a close family member and 9.09% (n=26) reported the death of an extended family member. Less than 1% of the respondents reported the death of a work colleague. When asked about the cause of death of the deceased, about two-thirds (67.23%, n=80) could not provide a specific cause while 11% described an acute illness and 22% described a chronic illness as the cause of death. This is shown in figure 5. Slightly more than half the (51.26%, n=61) respondents with COVID-19 associated symptoms reported knowing someone who died in their community in the preceding month. There was no statistically significant association between self-reporting of COVID-19 associated symptoms by respondents and reporting that they knew someone who died in their community during the month of Ramadan (p=0.207).

**Figure 4.**
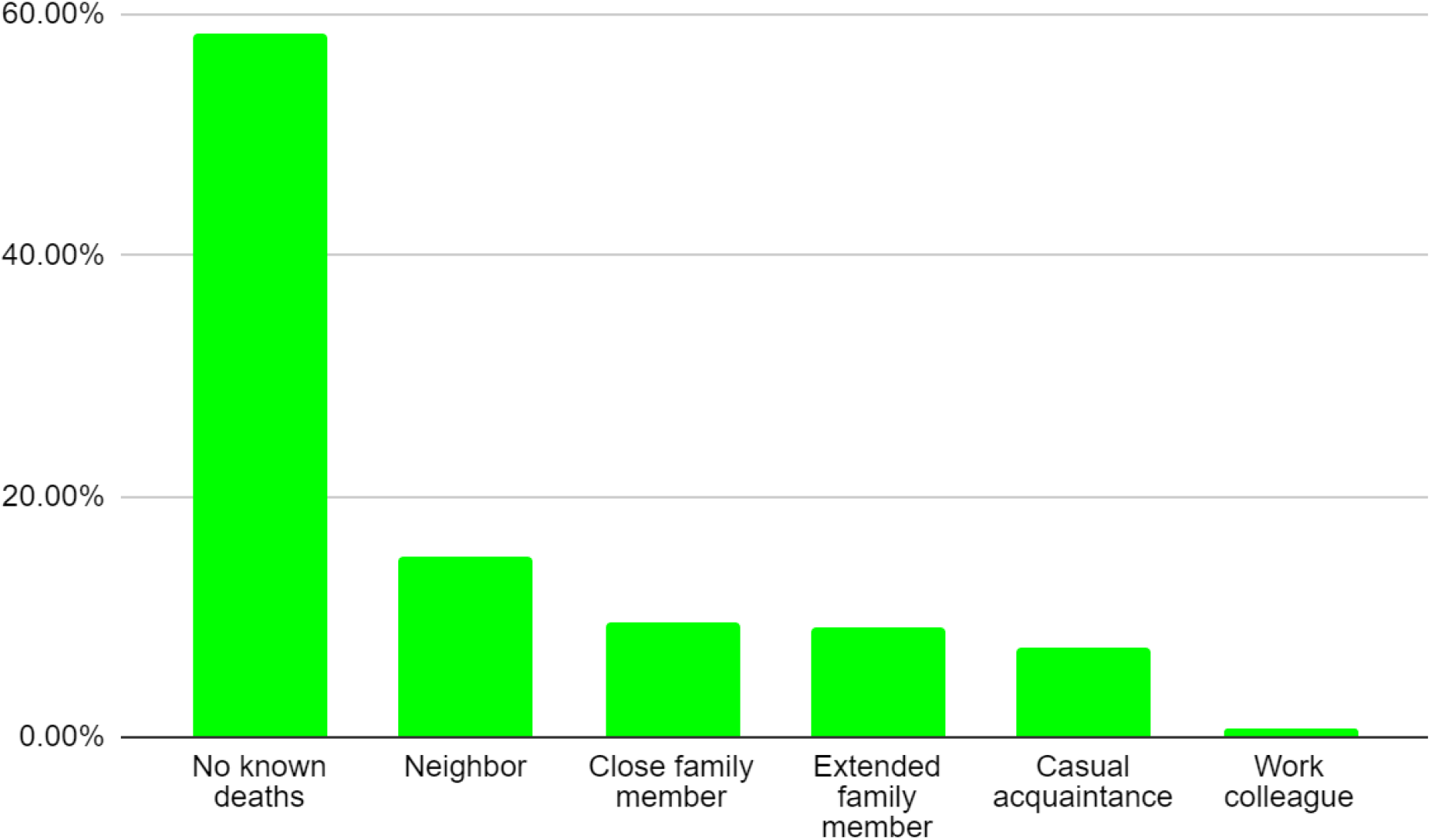
Community deaths reported by respondents

**Figure 5.**
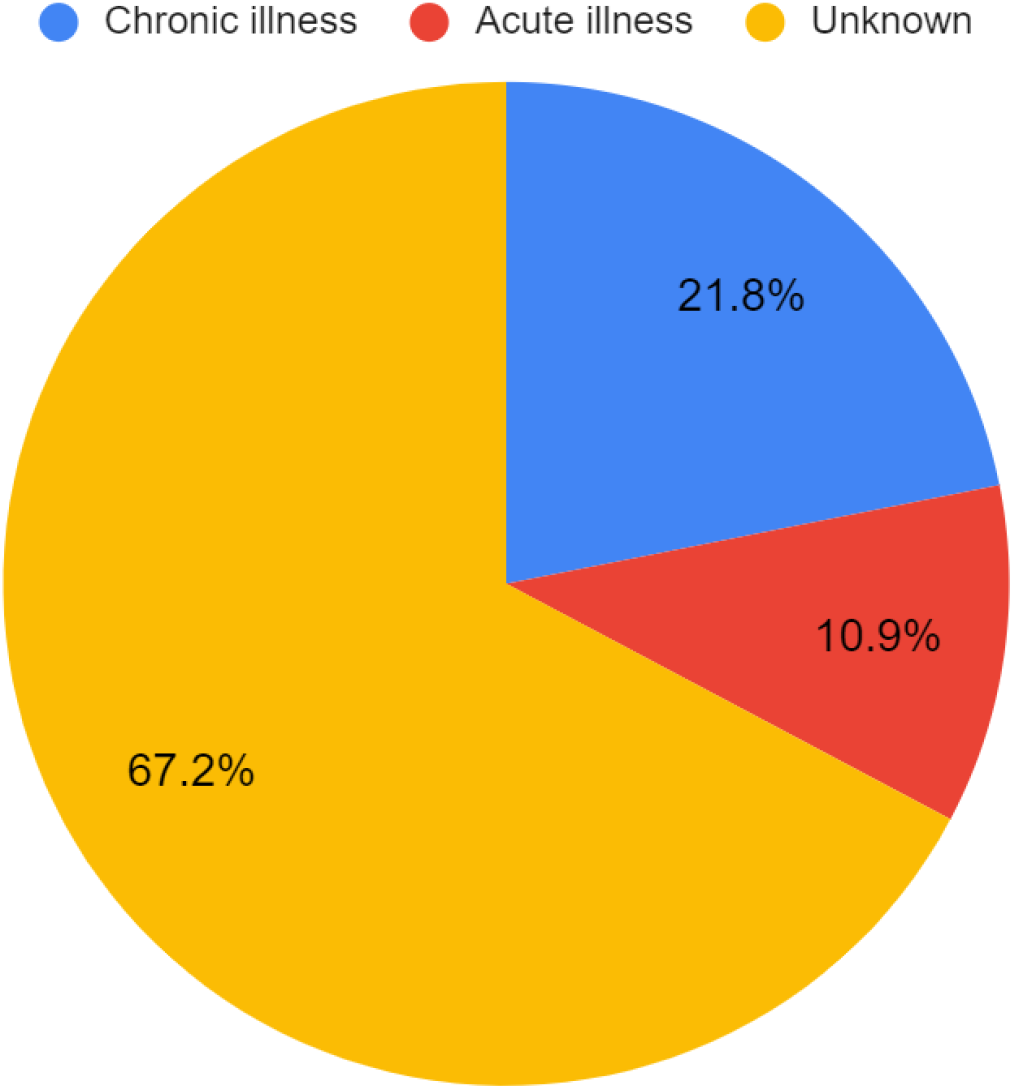
Cause of death reported by respondents

## Discussion

### Socioeconomic characteristics of our study population

This study surveyed the presence of COVID-19 associated symptoms and community deaths during the month of June 2020 amongst people in Kano city, Nigeria. Our survey cohort consisted of mainly young, educated and employed respondents. The Nigeria Demographic and Health Survey (DHS) 2018 reported 50% of adults aged 15-49 in Kano state as having no education and 12% having completed secondary education (Nigeria DHS 2018). However, Nigeria DHS 2018 did not include adult and Quranic education in its classification. Our study population had participants with higher levels of education even after accounting for this difference compared to the general population of Kano as indicated by the Nigeria DHS 2018 report. Nigeria DHS 2018 also reported 61% of adults aged 15-49 in Kano state as being employed at the time of the survey while we found 57% of our respondents to be employed. A hybrid online-offline COVID-19 knowledge, attitude and practice survey in Kano reported a study population with a mean age of 28.58, 55% male and 57% tertiary education (Habib, Dayyab et al. 2021). Our study population shares some general characteristics with those of the previous study such as a well-educated young population.

### COVID-19 associated symptoms

About half the respondents in our survey reported COVID-19 associated symptoms during the month of June, but only 1.4% of them were tested for COVID-19 despite a quarter of them seeing a health worker during the same period. We found a significant association between age and reporting COVID-19 associated symptoms where the odds of reporting symptoms increased with age. Age and male gender have been found to be strongly associated with positive COVID-19 tests in Nigeria (Elimian, Ochu et al. 2020). A community-based active surveillance for COVID-19 cases in Lagos found fever, cough and weakness to be the most common symptoms reported (Onasanya, Adebayo et al. 2020). However, none of these symptoms were found to have sufficient predictive value to make the diagnosis of COVID-19 when compared with SARS-CoV-2 PCR test. A retrospective data analysis of 36,496 patients tested for COVID-19 using PCR found those with loss of smell to be eight times more likely to be COVID-19 test positive compared to those that did not (Elimian, Ochu et al. 2020). The same study also reported fever, cough and loss of taste to be more likely in those who test positive for COVID-19. Elsewhere in the United States, loss of smell and loss of taste showed statistically significant association with positive COVID-19 PCR results (Yan, Faraji et al. 2020). A sudden increase in loss of smell in the setting of COVID-19 pandemic has been postulated to be indicative of presumed COVID-19 infection (Coelho, Kons et al. 2020).

Even though we found a statistically significant association between age and reporting COVID-19 associated symptoms in our study population, and age is an established risk factor for COVID-19 infection, we are left with the possibility that the symptoms reported by our survey respondents were caused by other causes and not COVID-19. Fever, cough and loss of appetite which were the most common COVID-19 associated symptoms seen in our survey can also be seen with the most commonly diagnosed infectious disease in Nigeria, malaria. A survey of 60 febrile patients aged 3-70 visiting a health center found 63% of them to have malaria (Ayorinde, Oyeyiga et al. 2016). Our survey was conducted in the month of June, when malaria incidence is low due to the prevailing dry and hot weather. This makes it more likely that the fever reported by 17% of respondents was due to causes other than malaria.

Another study of 179 adult patients with fever who received care at a hospital in Nigeria were found to be positive for Dengue virus infection in 69% of cases (Onoja, Adeniji et al. 2016). A systematic review of influenza epidemiology in Sub-Saharan Africa reported 10% of acute respiratory illnesses to be caused by influenza viruses (Gessner, Shindo et al. 2011). A study of 9,978 patients with severe acute respiratory infection and influenza-like illnesses in Uganda found 11.2% to be positive for Influenza A or B (Cummings, Bakamutumaho et al. 2016). These studies show that symptoms of fever, cough etc can be caused by a variety of pathogens including respiratory viruses other than SARS-CoV-2.

We saw decreasing odds of reporting COVID-19 associated symptoms among respondents with increasing levels of education compared to those with no education. However, the association between level of education and reporting of COVID-19 associated symptoms was not statistically significant. Similarly, level of education was found to have no significant association with good COVID-19 knowledge (Habib, Dayyab et al. 2021). Contradictory association with education level and COVID-19 positive test results has been noticed in other studies (Karout, Serwat et al. 2020).

Population level analysis in Nigeria found education and employment not to be associated with higher COVID-19 case mortality rate (Hassan, Hashim et al. 2020). We found respondents who had outdoor occupation to have twice the odds of reporting COVID-19 associated symptoms compared to those who are unemployed. It is conceivable that those in outdoor occupations such as traders and hawkers have higher odds of reporting symptoms since they come into contact with a large number of people in close proximity as they sell their wares in crowded outdoor markets. Interacting in close proximity has been shown to be a risk factor for transmitting SARS-CoV-2 (Chu, Akl et al. 2020).

Without access to COVID-19 diagnostic tests, we were limited in our scope by documenting COVID-19 associated symptoms and community deaths to paint a picture of the community during a peak of the pandemic in June 2020. Testing capacity for COVID-19 using PCR technique was limited in Nigeria as in most countries during the initial phase of the pandemic. In April 2020, there were only 9 diagnostic facilities nationwide, all in the public sector, that could conduct PCR testing for COVID-19 (Adesanya 2020). This increased to 62 facilities nationwide by August with the addition of private sector diagnostic laboratories that responded to the call from the government for more testing capacity (Adesanya 2020). Case definitions also varied as the pandemic evolved with epidemiological linkage as a requirement for COVID-19 testing initially but later dropped (Harapan, Itoh et al. 2020).

### Deaths

Media reports quoting grave diggers in Kano of unusually large numbers of deaths is partly what prompted us to conduct this COVID-19 associated symptom survey. Kano went under a 2-week lockdown in late April which was extended for a further 2 weeks in May (Adesanya 2020). There were reports of people defying the lockdown and religious services during this period across Nigeria (Ezeibe, Ilo et al. 2020). There were also reports of patients in COVID-19 isolation centers protesting their treatment conditions (Ezeibe, Ilo et al. 2020). A survey of communities showed that fear of COVID-19 kept people away from healthcare facilities and from seeking care when sick (Ahmed, Ajisola et al. 2020). Such a fear of health facilities in the middle of a pandemic combined with lockdowns and closure of routine healthcare could increase deaths in the community by preventing people from seeking healthcare not only for COVID-19, but also for other infectious diseases as well as exacerbation of chronic illnesses.

Our survey found slightly less than half the respondents (42%, n=119) reporting that they knew someone in the community who died during the month of June. About one-fifth of the respondents reported the death of a family member while 15% reported the death of a neighbor. No cause of death was known in two-thirds of cases while fever was the most frequently seen symptom in 29% of deaths.

Studies of excess mortality due to COVID-19 have compared mortality data from the study period with historic mortality data from the previous several years to arrive at estimates (Beaney, Clarke et al. 2020, Kung, Doppen et al. 2021, Modig, Ahlbom et al. 2021). Such a comparison is impossible in Nigeria where vital statistics registrations systems are weak and deaths in the community are often not registered (Sharma, Brown et al. 2017, Makinde, Odimegwu et al. 2020). Baseline and excess mortality for Nigeria is also not available from public databases such as The Human Mortality Database (Human Mortality Database accessed March 26, 2021) or from the World Mortality Dataset (Karlinsky and Kobak 2021). Nigeria had 1,915 confirmed deaths due to COVID-19 by March 1, 2021 (Our World in Data accessed March 5, 2021). This must be interpreted in light of a systematic literature review of death registration in Nigeria which reported that 10% of deaths were registered in 2017 (Makinde, Odimegwu et al. 2020). It is possible to estimate mortality rates from household surveys, but this requires careful planning and large sample sizes to ensure a representative sample of the target population is surveyed (Feehan, Mahy et al. 2017). Our sample size of 291 individuals and convenience sampling technique preclude any accurate estimation of mortality from the collected data. Notwithstanding this inability to estimate excess mortality due to COVID-19, our survey found a large proportion of respondents who reported deaths in their communities and a majority of these deaths were unexplained. It is noteworthy that chronic illnesses such as hypertension and diabetes were given as the cause in about a fifth of reported deaths even while fever was given as the most frequent symptom exhibited by the deceased individuals.

In addition to the limitations mentioned above such as lack of confirmatory lab test and baseline mortality data, we also acknowledge the small sample size as a limitation of our study. The sample population was small to perform sub-group analyses based on testing and symptom type. Additionally, over-reporting of symptoms due to prompts from the checklist could have been a possible source of bias leading to some over-estimated odds-ratios.

## Conclusions

Our survey of residents in urban Kano found half the respondents reporting at least one COVID-19 associated symptom in June 2020 and the odds of reporting symptoms increased with age. We did not find any statistically significant association between level of education and reporting of symptoms or gender and reporting of symptoms. Those with outdoor occupations were found to have twice the odds of reporting COVID-19 associated symptoms compared to the unemployed and this association was statistically significant. Slightly less than half the respondents also reported knowing someone in the community who died but no cause was given for two-thirds of those deaths. These findings provide circumstantial evidence of COVID-19 associated symptoms especially among the older population and deaths in an urban community in Kano. Lack of COVID-19 confirmatory laboratory tests and absence of baseline vital statistics precluded us from finding definitive evidence for or against COVID-19 infection and associated mortality.

## Supporting information

Raw survey data

## Data Availability

Data will be available under supplementary files.

## Acknowledgements

The authors wish to thank Mr. Tosin Williams of eHealth Africa for his assistance in creating the survey instrument.

## Notes

### Competing Interest Statement

The authors have declared no competing interest.

### Clinical Trial

This was not a clinical trial, but a survey of COVID-19 symptoms and mortality. Ethics committee approval was obtained from the local governing body.

### Funding Statement

No external funding was received for this project.

### Author Declarations

Ethics approval was obtained from Kano state government ministry of health (Approval number MOH/Off/797/T.I/2052). Informed verbal consent was obtained from all participants before data was collected from them.

